# A cohort study to evaluate the effect of combination Vitamin D, Magnesium and Vitamin B12 (DMB) on progression to severe outcome in older COVID-19 patients

**DOI:** 10.1101/2020.06.01.20112334

**Authors:** Chuen Wen Tan, Liam Pock Ho, Shirin Kalimuddin, Benjamin Pei Zhi Cherng, Yii Ean Teh, Siew Yee Thien, Hei Man Wong, Paul Jie Wen Tern, Manju Chandran, Jason Wai Mun Chay, Chandramouli Nagarajan, Rehena Sultana, Jenny Guek Hong Low, Heng Joo Ng

**Author notes:** both authors contributed equally (Chuen Wen Tan, Liam Pock Ho). **Corresponding author** A/Prof Heng Joo Ng, Address; Department of Hematology, Level 3, Academia, 20 College Road, Singapore 169856.

## Abstract

**Objective:** To determine the clinical outcomes of older COVID-19 patients who received DMB compared to those who did not. We hypothesized that fewer patients administered DMB would require oxygen therapy and/or intensive care support than those who did not.

**Methodology:** Cohort observational study of all consecutive hospitalized COVID-19 patients aged 50 and above in a tertiary academic hospital who received DMB compared to a recent cohort who did not. Patients were administered oral vitamin D3 1000 IU OD, magnesium 150mg OD and vitamin B12 500mcg OD (DMB) upon admission if they did not require oxygen therapy. Primary outcome was deterioration post-DMB administration leading to any form of oxygen therapy and/or intensive care support.

**Results:** Between 15 January and 15 April 2020, 43 consecutive COVID-19 patients aged ≥50 were identified. 17 patients received DMB and 26 patients did not. Baseline demographic characteristics between the two groups was significantly different in age. In univariate analysis, age and hypertension showed significant influence on outcome while DMB retained protective significance after adjusting for age or hypertension separately in multivariate analysis. Fewer DMB patients than controls required initiation of oxygen therapy during their hospitalization (17.6% vs 61.5%, P=0.006). DMB exposure was associated with odds ratios of 0.13 (95% CI: 0.03 – 0.59) and 0.20 (95% CI: 0.04 – 0.93) for oxygen therapy and/or intensive care support on univariate and multivariate analyses respectively.

**Conclusions:** DMB combination in older COVID-19 patients was associated with a significant reduction in proportion of patients with clinical deterioration requiring oxygen support and/or intensive care support. This study supports further larger randomized control trials to ascertain the full benefit of DMB in ameliorating COVID-19 severity.

## Introduction

The COVID-19 pandemic which began in late 2019 has raged across the globe with more than four million infections and 300,000 deaths recorded to date. A broad theme of immune hyper-inflammation has emerged as a key determinant of patient outcome with uncontrolled immune response postulated as a pathophysiologic factor in disease severity.^1^ Intuitively, immunomodulation becomes an attractive potential treatment strategy. Besides lung involvement, gastrointestinal symptoms are frequent and carry a worse prognosis.^2^ COVID-19 is therefore a multi-organ phenomenon and it is becoming evident that appropriate systemic inflammatory control is necessary for overall survival benefit. Patient factors such as age>50, hypertension, diabetes and coronary artery disease, are also known associations with increased severity and death.^3,4^

Much of the current therapeutic effort is targeted at viral elimination instead of pre-emptively modulating hyper-inflammation. A number of immunomodulatory agents may serve the latter role. Vitamin D for instance, has a protective effect against respiratory tract infection,^5^ while magnesium enhances Vitamin D function besides being a vasodilator, anti-thrombotic and bronchodilator.^6,7^ Lastly, vitamin B12 is an important modulator of gut microbiota.^8^ Importantly, these compounds are generally safe and well-tolerated by patients. A short course of these three supplements (DMB) upon diagnosis of COVID-19 could potentially exert synergistic effects to modulate host immune response, ameliorate COVID-19 severity and reduce adverse outcomes. This study was conducted to evaluate the potential efficacy of DMB on progression of COVID-19 to severe disease.

## Methods

### Study Design

This study was approved by our institutional ethics committee with waiver of consent granted (Ref No:2020/2344). We included all consecutive COVID-19 patients aged 50 years and above admitted to Singapore General Hospital, a tertiary academic hospital, between 15 January and 15 April 2020. Diagnosis required a positive SARS-CoV-2 PCR from nasopharyngeal or throat swab. Primary outcome was defined as the requirement of any form of oxygen therapy, and/or intensive care unit (ICU) support. As the COVID-19 situation evolved, we decided from 6 April 2020, to start DMB on all COVID-19 patients above 50 years old if they did not require oxygen therapy. Similar patients admitted before this date did not receive DMB and therefore served as the control population. Therapy comprised a single daily oral dose of vitamin D3 1000 IU, magnesium 150mg and vitamin B12 500mcg for up to 14 days. DMB could be stopped if a patient subsequently deteriorated or deemed to have recovered based on symptom resolution and 2 consecutive negative SARS CoV-2 RT-PCR respiratory samples.

### Data collection

Clinical and laboratory data were collected from electronic health records in a standardized form with two investigators independently reviewing the data for accuracy.

### Statistical Analysis

Primary outcome requiring oxygen therapy was treated as binary data with categories *‘yes’* or *‘no’*. Demographic and clinical characteristics were summarized with respect to intervention and control group. Continuous variables were expressed as mean/standard deviation (SD) if normal distribution, or median (interquartile range) if non-normal distribution, and categorical data was expressed as number counts and percentages as appropriate. Univariate and multivariable binary logistic regression was performed to find associated risk factors for primary outcome. Quantitative association from logistic regression was expressed as odds ratio (OR) with 95% confidence interval (95%CI). All tests were two sided and p-value <0.05 was considered as statistical significance. Statistical software SPSS version 25 (IBM, USA) was used for analysis.

### Results

43 consecutive patients were identified with 17 patients in the DMB arm and 26 patients in the control arm. Baseline demographic and clinical characteristics was significantly different for age between the two groups. (Table 1) In the treatment arm, most patients received DMB within the first day of hospitalization with a median duration of therapy of 5 days (interquartile range of 4 to 7 days). Significantly fewer patients receiving DMB required subsequent oxygen therapy compared to controls (3/17 vs 16/26, P=0.006) (Table 1 & Figure 1). On univariate analysis, increasing age and hypertension demonstrated significantly higher odds ratio for oxygen therapy, while exposure to DMB therapy was associated with a significantly improved odds ratio. (Table 2) Multivariate analysis showed that DMB remained a significant protective factor against clinical deterioration after adjusting for age or hypertension separately.

**Table 1:** Baseline demographic and clinical characteristics and outcomes of the patients given DMB therapy and control patients. DMB = Vitamin D, Magnesium, Vitamin B12, SD = Standard Deviation, IQR = Interquartile Range, CXR = chest x-ray, ICU = intensive care unit.

|  | DMB (N = 17) | Control (N = 26) | P-value |
| --- | --- | --- | --- |
| **Baseline characteristics** |  |  |  |
| Age, years, mean (SD) | 58.4 (7.0) | 64.1 (7.9) | **0.021** |
| Male, n (%) | 11 (64.7) | 15 (57.7) | 0.755 |
| Female, n (%) | 6 (35.3) | 11 (42.3) | 0.755 |
| Comorbidities, n (%) | 8 (47.0) | 20 (76.9) | 0.057 |
| Diabetes mellitus | 0 (0.0) | 6 (23.1) | 0.066 |
| Hypertension | 6 (35.3) | 18 (69.2) | 0.058 |
| Hyperlipidemia | 5 (29.4) | 15 (57.7) | 0.118 |
| Cardiovascular Disease | 1 (6.0) | 6 (23.0) | 0.376 |
| Asthma / COPD | 2 (11.7) | 2 (7.7) | 1.000 |
| Stroke | 0 (0.0) | 2 (7.7) | 0.511 |
| **Clinical features** |  |  |  |
| Normal CXR on admission, n (%) | 7 (41.2) | 7 (26.9) | 0.507 |
| Time from onset of symptoms to admission, days, median (IQR) | 7 (1-9) | 5 (3-8) | 0.455 |
| Time from onset of symptom to initiation of therapy, days, median (IQR) | 7 (4-10) |  |  |
| Time from admission to initiation of therapy, days, median (IQR) | 1 (0-1) |  |  |
| Duration of therapy, days, median (IQR) | 5 (4-7) |  |  |
| **Outcome** |  |  |  |
| Requiring any form of oxygen therapy (including ICU support), n (%) | 3 (17.6) | 16 (61.5) | **0.006** |
| - Requiring oxygen therapy (but not ICU support), n (%) | 2 (11.8) | 8 (30.8) |  |
| - Requiring ICU support, n (%) | 1 (5.9) | 8 (30.8) |  |

**Table 2:**
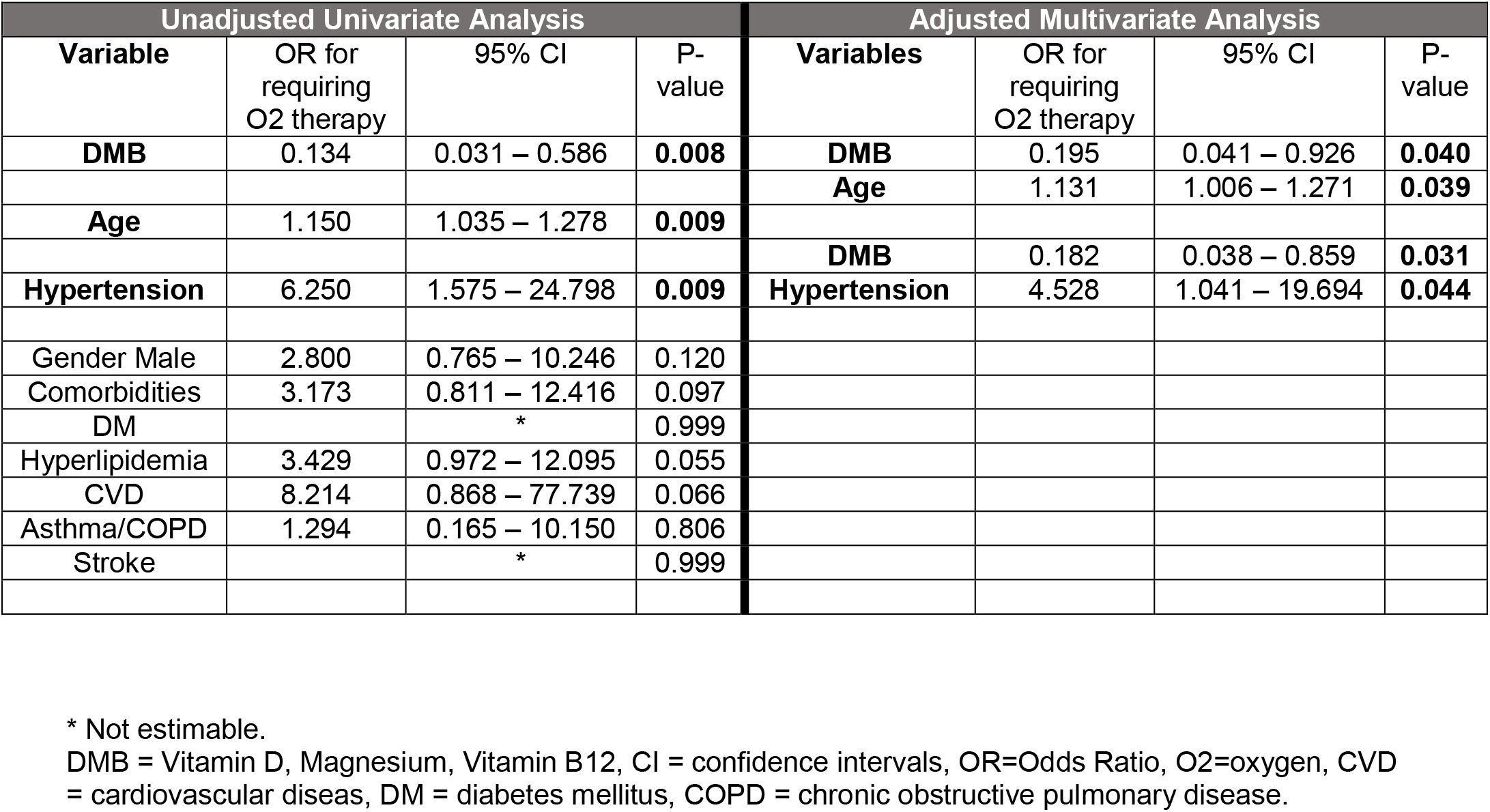
Univariate and multivariate analyses of odds ratio in developing primary outcome requiring oxygen therapy for Clinical variables and DMB therapy. * Not estimable. DMB = Vitamin D, Magnesium, Vitamin B12, CI = confidence intervals, OR=Odds Ratio, O2=oxygen, CVD = cardiovascular diseas, DM = diabetes mellitus, COPD = chronic obstructive pulmonary disease.

**Figure 1:**
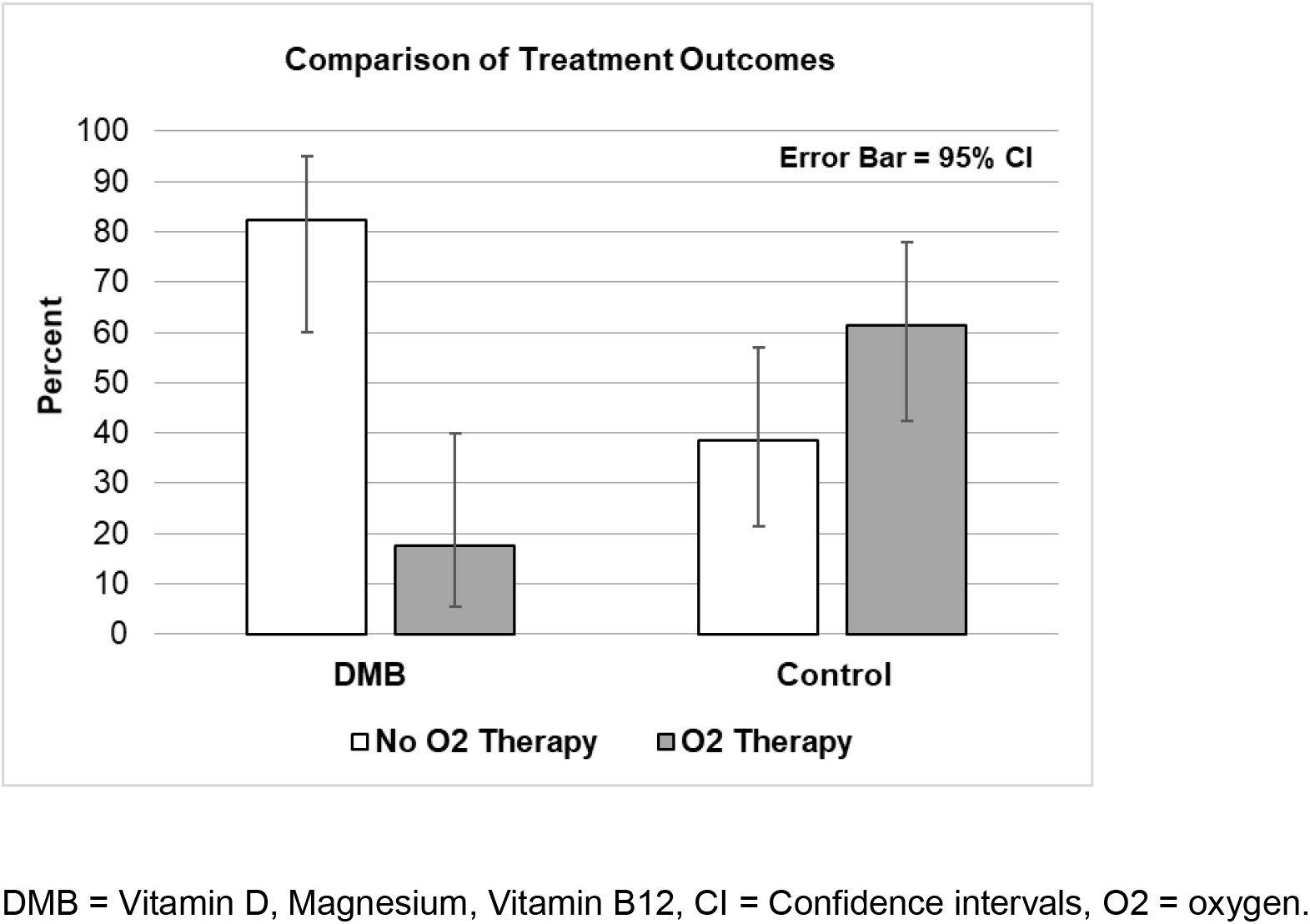
Treatment outcomes in terms of requirement of oxygen therapy of patients who received DMB and control patients. DMB = Vitamin D, Magnesium, Vitamin B12, CI = Confidence intervals, O2 = oxygen.

All patients who needed supplemental oxygen therapy in the control group also required further ICU support. Of the three patients who required oxygen therapy in the DMB group, two patients were started on oxygen therapy within 24 hours from the initiation of DMB. The third patient required supplemental oxygen therapy after 3 days of DMB but did not require ICU support. Among 9 patients given DMB within the first week of onset of symptoms, only one patient required oxygen therapy. This patient was one of the two cases which deteriorated within 24 hours of DMB initiation.

Of note, there were no side effects or adverse events directly attributable to DMB.

### Discussion

COVID-19 is now understood to be potentially life-threatening in up to 20 percent of patients. As the world awaits an effective vaccine, the effectiveness of various antivirals are largely muted by lack of survival benefit. Targeted therapies against cytokines and anti-thrombotic agents may only address the terminal events in severe cases with limited benefits. At the point of giving DMB to our older patients, it became obvious that pre-emptive down-regulation of hyper-inflammation with relatively safe agents was an attractive alternate strategy. This combination was chosen based on substantial, albeit indirect evidence of their role in tempering the inflammatory response to viral infections. Vitamin D, through its effect on NFkB and other pathways, can attenuate various proinflammatory cytokines^9^ mediating the uncontrolled cytokine storm seen in severe COVID-19 with deficiency associated with severe COVID-19.^10^ Magnesium is critical in the synthesis and activation of vitamin D, acting as a cofactor in many of the enzymes involved in vitamin D metabolism.^11^ Vitamin B12 is essential in supporting a healthy gut microbiome which has an important role in the development and function of both innate and adaptive immune systems.^12^ This could be pivotal in preventing excessive immune reaction^13^ especially in COVID-19 patients with microbiota dysbiosis which were associated with severe disease.^14^

Our results provide early positive evidence of an immune-modulatory approach to ameliorating severe outcome in COVID-19. DMB treated patients were significantly less likely to require oxygen therapy compared to controls. Among three DMB treated patients with clinical deterioration, two likely deteriorated within 24 hours from their underlying infection but were included on an intention-to-treat analysis. Had they been excluded on the basis of inadequate time of DMB exposure, the demonstrated benefits would have been more profound. The last case who deteriorated was started on DMB after 7 days from onset of symptoms. To benefit from its pre-emptive effects, patients may need to be started earlier in the infection course. The ease of administration of DMB would also allow for early initiation in primary care setting at first onset of symptoms, or as prophylaxis among high risk contacts during outbreaks in identified clusters.

As all agents in this combination are readily available, safe and inexpensive, DMB can benefit a large swath of the world population especially in economically-challenged countries with limited or late access to vaccines and other therapies. DMB may also exhibit a generic efficacy against other viral infections with similar pathological mechanism.

This study was conducted under difficult dynamic circumstances and is thus limited by the small sample size, and the lack of systematic biologic measures to support its findings. It is however a proof-of-principle effort with promising results. Our findings would need to be further validated in a well-designed randomized study.

## Data Availability

The data for this manuscript is available from the corresponding author.

## Author contributions

CWT, LPH, SK, JGL and HJN co-wrote the manuscript. CWT, LPH, JWMC, MC, SK, JGL and HJN were involved in the design of the study. BPZC, YET, SYT, HMW, PJWT, JGL and SK conducted the pilot study. CWT, LPH, CN and RG were involved in data analysis. LPH formulated the supplement combination. All authors have read and agreed with the manuscript.

## Conflict of Interest

All authors have nothing to declare.

## Acknowledgments

The authors are grateful for the continued support from SingHealth and Singapore General Hospital. We are also grateful to all doctors and nurses who cared for the patients in the isolation wards, SGH.

## Notes

### Competing Interest Statement

The authors have declared no competing interest.

### Clinical Trial

It is registered under ISRCTN15324611

### Funding Statement

Not a funded study.

### Author Declarations

This study was approved by our institutional ethics committee with waiver of consent granted (Ref No:2020/2344).

